# Prevalence and Factors Associated with Behavioural Problems in 5-year-old Children Born with Cleft Lip and/or Palate from the Cleft Collective

**DOI:** 10.1101/2021.08.04.21261594

**Authors:** Samantha Berman, Gemma C Sharp, Sarah J Lewis, Rachel Blakey, Amy Davies, Kerry Humphries, Yvonne Wren, Jonathan R Sandy, Evie Stergiakouli

**Author notes:** Corresponding Author: Dr Evie Stergiakouli.

## Abstract

**Objectives:** We determined the prevalence of behavioural problems in 5-year-old children born with Cleft Lip and/or Palate (CL/P) and compared it to the prevalence in general population samples. We also identified risk factors for behavioural problems in children with CL/P.

**Design:** Observational study using questionnaire data from the Cleft Collective (CC) 5-Year-Old cohort study and three general population samples.

**Main Outcome Measure:** The Strengths and Difficulties Questionnaire (SDQ) was used to measure behavioural problems.

**Participants:** A total of 340 children born with CL/P whose mothers had completed the SDQ when their child was 5 years old. Published estimates from three large cohorts were used to approximate general population SDQ scores in the UK and these were used as comparison groups; Millennium Cohort Study (MCS) (n=12,511), Office of National Statistics (ONS) normative school-age SDQ data (n=5,855) and the Avon Longitudinal Study of Parents and Children (ALSPAC) (n=9,386).

**Results:** An estimated 14.3% of 5-year-old children born with CL/P experienced behavioural problems. There was strong evidence to suggest children with CL/P were more likely to experience difficulties than children in the general population, as measured by SDQ total difficulties scores from all three population cohorts: MCS (OR = 2.07 [95% CI = 1.50-2.85]; P<.001), ONS Norms (OR = 1.53 [95% CI = 1.12-2.11]; P=.008), and ALSPAC (OR = 2.37 [95% CI = 1.72-3.27]; P<.001). The odds of hyperactivity, emotional, prosocial and peer problems were increased among children in the Cleft Collective compared with children in the Millennium Cohort Study. Odds ratios and 95% confidence intervals indicated that children in the Cleft Collective were nearly twice as likely as those in the MCS (OR = 1.91 [95% CI = 1.38-2.65]; P<.001) and three times those in ALSPAC (OR = 3.20 [95% CI = 2.29-4.47]; P<.001) to experience emotional difficulties. The odds of emotional difficulties were higher in boys than girls. Maternal smoking, marital status, younger maternal age at conception, lower maternal education, receiving income support, and measures of poor maternal and familial health showed some evidence of association with behavioural problems in 5-year-old children born with CL/P.

**Conclusions:** Our findings suggest elevated levels of behavioural problems in children born with CL/P, particularly emotional difficulties in boys, compared to the general population and indicate several factors associated observationally with these difficulties.

## Introduction

Cleft of the lip and/or palate (CL/P) is one of the most common congenital anomalies affecting around 1000 babies born in the United Kingdom (UK) every year (*Cleft Registry and Audit Network (CRANE) Database 2018 Annual Report*.). Worldwide, approximately 1 in 700 children are born with an orofacial cleft. Children can have a cleft of the lip (CL), cleft of the palate (CP), or a combination of the two (cleft lip and palate [CLP]). While prognosis is generally good, children born with CL/P undergo several repair operations, show physical signs of scarring and often experience breathing, feeding, speech, dental and hearing problems (M. C. Endriga & Kapp-Simon, 1999).

In recent years, research has focused on the wider implications of cleft, particularly the psychological and social impacts, challenging the traditional concept of cleft as a cosmetic and functional condition (Al-Namankany & Alhubaishi, 2018; Broder & Strauss, 1989; Feragen & Stock, 2016; Thomas, Turner, Rumsey, Dowell, & Sandy, 1997). Speech and language difficulties, poor self-image, and negative social experiences, compounded by the stress of ongoing medical treatment, have been thought to increase risk of behavioural problems in children born with CL/P (Lockhart, 2003; Snyder, Bilboul, & Pope, 2005; Strauss et al., 2007).

Psychological functioning and behaviour difficulties in children are strong predictors of future mental health (Sosu & Schmidt, 2016). Studies have linked difficulties in childhood to academic underachievement, psychosomatic disorders, unemployment, and an overall reduced quality of life (Fitzsimons et al., 2018; O. Hunt, Burden, Hepper, & Johnston, 2005; Orlagh Hunt, Burden, Hepper, Stevenson, & Johnston, 2007; Sosu & Schmidt, 2016; Stock, Feragen, & Rumsey, 2015; Thamilselvan, Kumar, Murthy, Sharma, & Kumar, 2015) As an early diagnosis of psychiatric disorders is known to proffer better health outcomes and diminish economic and societal costs, there is an impetus to identify children who may be at an elevated risk for developing difficulties (Sosu & Schmidt, 2016).

Children born with CL/P typically present with subclinical mental health and wellbeing needs (Stock & Feragen, 2016), which are negatively impacted when the cleft is combined with additional conditions and stresses(Feragen & Stock, 2014). Evidence from longitudinal and cross-sectional studies suggests that children born with CL/P present with behavioural problems at ages 5 and 10 years more frequently than children without CL/P in the general population (Pinckston, Dalton, Farrar, & Hotton, 2020; Waylen et al., 2017). However, large-scale individual studies on clinical populations are rare, and more research is required to ascertain the prevalence of mental health problems and to examine the possible unmet needs of this group (Berger & Dalton, 2011; Feragen, Særvold, Aukner, & Stock, 2017; Klassen et al., 2012; Thomas et al., 1997).

There is evidence to suggest that children born with CL/P struggle with difficulties related to conduct (Richmen & Millard, 1997), social skills (Brand et al., 2009; Murray et al., 2010; Richmen & Millard, 1997), self-regulation (M. C. Endriga, Jordan, & Speltz, 2003), and hyperactivity (Waylen et al., 2017). Such behavioural problems, along with cleft-related functional challenges and aesthetic differences, have been associated with mother-to-infant bonding issues (Marya C. Endriga & Speltz, 1997; Maris, Endriga, Speltz, Jones, & DeKlyen, 2000), teasing (Lorot-Marchand et al., 2015), stigma (Alansari, Bedos, & Allison, 2014; Strauss et al., 2007) social isolation (O. Hunt et al., 2005; Murray et al., 2010; Noar, 1992; Ramstad, Ottem, & Shaw, 1995) and psychological ajustement issues (Feragen et al., 2017; Millard & Richman, 2001).

Sociodemographic factors and maternal characteristics are known to affect mental health and behavioral outcomes in children (Kovess-Masfety et al., 2016). Studies based on large cohorts, such as the Avon Longitudinal Study of Parents and Children (ALSPAC) and the Millennium Cohort Study (MCS), have contributed to our understanding of the relative predictive power of parental health and sociodemographic variables on childhood behavioral difficulties in the general UK population (Borrell-Porta M, 2017; Washbrook). These associations have not been well explored in a cleft population.

The aim of this study was to estimate the prevalence of behavioural problems in 5-year-old children born with CL/P and compare it to that estimated in children of similar age from the general population. In addition, we tested if maternal, familial and cleft-related factors are associated with behavioural problems in children born with CL/P.

## Methods

### Study Sample

The current study used data from the Cleft Collective (CC) 5-Year-Old Cohort. The Cleft Collective is an ongoing UK-wide longitudinal study, comprising a Birth Cohort and a 5-Year-Old Cohort, established to investigate the causes, best treatments, and the psychological impact of cleft, on affected individuals and their families. Data from the Cleft Collective is available for clinical and academic communities to access and use to address a range of cleft related research questions. More information on the study and how to access the dataset is available at http://www.bristol.ac.uk/cleft-collective/professionals/access/. A detailed description of recruitment procedures and the study’s development can be found elsewhere (Stock & Feragen, 2016). The Cleft Collective Cohort Studies have been approved by the South West Central Bristol NRES ethics Committee (REC 13/SW/0064). This study was approved by the University of Bristol Faculty of Health Sciences Student Research Ethics Committee (HSSREC).

### Strengths and Difficulties Questionnaire

The Strengths and Difficulties Questionnaire (SDQ) was given to all parents/guardians in the Cleft Collective Five-Year-Old Cohort as part of a larger baseline questionnaire. SDQ is one of the most widely used screening tools, adopted globally as a measure of mental health (A. Goodman & Goodman, 2009). The 25-item survey assesses a child’s mental health across five subscales: emotional symptoms, conduct problems, hyperactivity, peer-related behavioral problems, and prosocial behavior (R. Goodman, 1997). The instruments’ validity and reliability, including internal consistency, test-retest reliability, and inter-rater agreement have been established, and it has been shown to be strongly correlated with other tools (R. Goodman, 1997; R. Goodman, Ford, Simmons, Gatward, & Meltzer, 2000; Stone, Otten, Engels, Vermulst, & Janssens, 2010). Previous studies have shown that although SDQ subscales vary, parent-reported total difficulties scores have good internal reliability (Van Roy, Grøholt, Heyerdahl, & Clench-Aas, 2006).

In this study, parents completed the SDQ on behalf of the study child. Parents answered questions relating to their child’s behavior using a three-point rating scale: not true (=0), somewhat true (=1) and certainly true (=2). Each subscale, comprised of five questions, was then scored on a scale of 0-10. Anchor points were reversed for the four scales measuring difficulties (emotional, conduct, hyperactivity, and peer problems) so that high scores in these categories represent greater difficulties. Total difficulties scores were calculated by summing scores from the four subscales; conduct problems, hyperactivity, emotional and peer problems. The prosocial scale was scored as is, with high scores in this domain indicating fewer difficulties. For the purposes of the current study, we used reports from mothers only (i.e. not partners or other guardians) for consistency and because these were more complete.

Given a skewed distribution of the total difficulties score, and each of the subscale scores, SDQ scores were dichotomized based on validated cut-points. Scores above subscale-specific thresholds (≥ 4 for conduct, ≥ 5 for emotional, ≥ 7 for hyperactivity, ≥ 4 for peer problems, ≥ 6 for prosocial) were designated as “cases”, signaling being at high risk of these behavioural problems, and scores below were classified as or “non-cases”(R. Goodman, 1997; R. Goodman et al., 2000). The overall total difficulties score ≥ 17, is henceforth used to indicate an individual at high risk of behavioural problems.

### Maternal, familial and cleft-related factors tested for association with behavioural problems

From the Cleft Collective Five-Year-Old Cohort baseline questionnaire completed by the mothers, the following variables were generated and tested for association with behavioural problems: Child’s sex (male, female; obtained from parental-report and medical notes, where this information was not available sex was assumed by the Cleft Collective operations team based on name and confirmed with cleft teams where sex could not be determined); Cleft type (CL, CP, CLP; derived from multiple sources including parental-report and medical notes); Syndromic cleft (no, yes confirmed or suspected; derived from multiple sources including parental-report and medical notes); Maternal age at conception (dichotomized as >24 years, ≤24 years; self-reported); Mother’s ethnicity (white, Black/Asian/minority ethnic (BAME); self reported); Mother’s highest educational qualification (≥ university degree, no degree; self-reported); Derived household annual income (≥ £20,000, <£20,000; self-reported, mothers and partners were asked about their annual income separately. If they lived together, incomes were summed together. If mother lived on their own, only mother’s income was reported); Mother receives income support (no, yes; self-reported); Number of biological children prior to the study child (parity) (0, ≥ 1; self-reported); Mother currently smokes cigarettes (no, yes; self-reported); Mother’s marital status (married/partnered, single/divorced/widowed; self-reported); Mother currently drinks alcohol (no, yes; self-reported); Perceived Stress Scale (PSS) (<50%, ≥ 50%-scores from the PSS were dichotomized on the 50th percentile (%) mark for the study cohort (Cohen, 1988)); PedsQL™ Family Impact Module (FIM) total, family functioning, maternal health-related quality of life (HRQOL) (<50%, ≥ 50%-scores from the PedsQL™ FIM were dichotomized on the 50th percentile (%) mark for the study cohort (James W. Varni, Sherman, Burwinkle, Dickinson, & Dixon, 2004)).

### General Population Estimates (control samples)

Published estimates from three large cohorts were used to approximate general population SDQ scores in the UK. SDQ scores differ by age, gender, and context specific factors, necessitating careful interpretations of normative data averages used to make cross-cohort comparisons (Croft, Stride, Maughan, & Rowe, 2015; R. Goodman, 2001). The Millennium Cohort Study (MCS) was used as the primary control group in this study, as the age of participants and recruitment timeline most closely match the CC ((CLS), 2018). The MCS third survey sweep provided SDQ estimates from a UK wide sample of 5-year-olds (n=12,511) ((CLS), 2018), estimates are published https://cls.ucl.ac.uk/cls-studies/millennium-cohort-study/mcs-age-5-sweep/. The study includes children born between September 2000-January 2002, and intentionally over-sampled children of ethnic minority backgrounds as well as those living in resource poor areas, who may otherwise have lower response rates, in order to ensure a representative sample ((CLS), 2018). The MCS aligns with our study sample in terms of age, timing, and geographical representation, although we were unable to obtain gender-specific estimates for MCS.

In addition, normative (Norms) school-age SDQ data from the UK’s Office of National Statistics (ONS), split by gender-and age-band are published (Sdqinfo.com, 2016) and are widely cited in the literature (Meltzer, Gatward, Goodman, & Ford, 2003; Waylen et al., 2017). Estimates were established from a large national survey carried out by the ONS in 1999 and include a representative sample of (n=5,855) parent-reported SDQ data for children aged 5-10. More information about the study sample can be found elsewhere (Meltzer et al., 2003). These general population estimates represent the most widely used British “norms”, but their comparability with our study sample is limited by large age-bands and temporal differences.

Finally, we calculated mean SDQ scores in The Avon Longitudinal Study of Parents and Children (ALSPAC), which is a transgenerational prospective study which recruited pregnant women resident in Avon, UK with delivery dates between 1^st^ April 1991-31^st^ December 1992 (Boyd et al., 2013; Fraser et al., 2013). Mothers (n=9,386) from the ALSPAC study completed the SDQ on their children between the ages of 3.7 and 5.4 (M=4.0 SD=0.12). This cohort provided general population estimates from a large sample and offered gender-specific estimates. Though there was a slight age difference between the samples, the age bracket for this control was small and overlapped the study cohort. The comparability of this sample with our cohort was limited by generational and geographical differences. The study website contains details of all the data that is available through a fully searchable data dictionary http://www.bristol.ac.uk/alspac/researchers/our-data/ and variable search tool. More information can be found online http://www.bristol.ac.uk/alspac/researchers/participants/.

Ethical approval for the ALSPAC study was obtained from the ALSPAC Law and Ethics committee and local research ethics committees. Published data on SDQ from the Millennium Birth Cohort and the UK norms was used so separate ethics approval was not sought for these studies.

### Statistical Analysis

SDQ scores were calculated for all participants in Stata, using the scoring guide published (Sdqinfo.com, 2016). Parent-reported SDQ scores from the Cleft Collective were analyzed as both continuous and categorical variables. First, Stata was used to calculate mean scores and standard deviations (SD) for each SDQ subscale.

We calculated odds ratios of children in the Cleft Collective having behavioural problems compared to those in the general population cohorts. Odds ratios and 95% confidence intervals were calculated to compare the proportion of high total difficulties score “cases” in the Cleft Collective to those in each of the three cohorts. Analyses were repeated for “cases” across each SDQ subscale. All analyses were repeated stratified by gender.

Logistic regression was used to assess associations between dichotomized SDQ scores (dependent variable) and maternal, familial and cleft-related (independent) variables. In each analysis, binary SDQ scores defined the dependent variable, and the maternal factors were used as binary independent variables, except cleft-type which was treated as a nominal categorical variable. We reported the odds ratios (OR) with 95% confidence intervals and gender-adjusted odds ratios for each of the variables across subscales.

## Results

### Sample Description

Eight hundred and ninety eight mothers had been recruited to the Cleft Collective 5-Year-Old Cohort at the point of analysis, data were available for n=340 5-year-olds with CL/P at recruitment resulting in a response rate of 37.9%. Table 1 summarizes the maternal and sociodemographic characteristics of the Cleft Collective study sample.

**Table 1:** Overview of Sociodemographic and Maternal Characteristics and Behavioural Problems across the Cleft Collective

| Cleft Collective<br>(Mothers returned baseline questionnaire (BLQ) at age 5) |  |  |
| --- | --- | --- |
| Variable | n(%) Total | SDQ Total Difficulties<br>(≥17)<br>n(%) Cases |
| **Mothers returned (BLQ)** |  |  |
| Yes | 340 | 46(13.53) |
| **Gender** |  |  |
| Male | 174(51.94) | 30(18.07) |
| Female | 161(38.06) | 16(10.39) |
| **Cleft type** |  |  |
| Cleft Lip | 41(24.12) | 6(15.00) |
| Cleft Palate | 59(34.71) | 8(14.29) |
| Cleft Lip and Palate | 70(41.18) | 11(16.42) |
| **Syndrome** |  |  |
| No | 84(89.36) | 10(12.50) |
| Yes | 10(10.64) | <5 |
| **Maternal Age at conception ≤24** |  |  |
| No | 270(83.33) | 26(10.04) |
| Yes | 54(16.67) | 15(29.41) |
| **Maternal Ethnicity** |  |  |
| White | 303(92.66) | 42(14.38) |
| BAME | 24(7.34) | <5 |
| **Maternal Education (≥Degree or equivalent)** |  |  |
| Yes | 149(46.71) | 13(8.97) |
| No | 170(53.29) | 30(18.75) |
| **Household Income ≥£20,000 annual** |  |  |
| Yes | 223(71.70) | 29(13.55) |
| No | 88(28.30) | 13(15.29) |
| **Mother Receives Income Support** |  |  |
| No | 302(93.50) | 36(12.41) |
| Yes | 21(6.50) | 9(47.37) |
| **Parity** |  |  |
| 0 | 174(53.21) | 27(15.98) |
| ≥1 | 153(46.79) | 16(11.03) |
| **Mother Smokes** |  |  |
| No | 116(73.89) | 10(9.52) |
| Yes | 41(26.11) | 13(31.71) |
| **Marital Status** |  |  |
| Married/Partnered | 288(86.49) | 33(11.96) |
| Single/Divorced | 45(13.51) | 13(30.23) |
| **Mother Consumes Alcohol** |  |  |
| No | 106(31.45) | 14(13.73) |
| Yes | 231(68.55) | 31(14.03) |
| **Maternal PSS≥50%** |  |  |
| No | 156(49.21) | <5 |
| Yes | 161(50.79) | 39(25.49) |
| **PEDSQL (Total score ≥50%)** |  |  |
| No | 166(49.85) | 9(5.56) |
| Yes | 167(50.15) | 37(23.42) |
| **PEDSQL (Family Functioning ≥50%)** |  |  |
| No | 196(58.68) | 13(6.84) |
| Yes | 138(41.32) | 33(25.19) |
| **PEDSQL (Maternal HRQOL ≥50%)** |  |  |
| No | 146(50.00) | 6(4.23) |
| Yes | 146(50.00) | 34(24.82) |

*\*BLQ=Baseline Questionnaire from Cleft Collective 5-Year-Old Cohort; ‘Cases’ derived from SDQ=Strengths and Difficulties Questionnaire total difficulties scores (Goodman, 1997); cell counts of less than 5 cannot be published for disclosure purposes.*

### Prevalence of behavioural problems among the Cleft Collective five-year-old cohort

Overall 14.3% of the n=340 children from the CC study sample were at high risk of mental health problems. Table 2 shows the frequency (n) and percentage (%) of “cases” in the Cleft Collective. Among boys 14.8% were classed as cases; among girls 9.1% in the cohort. The hyperactivity and prosocial subscales showed the greatest gender disparities (hyperactivity difficulties: 23.2% of boys and 14.3% of girls, prosocial difficulties: 23.4% of boys and 15.8% of girls).

**Table 2a & b:**
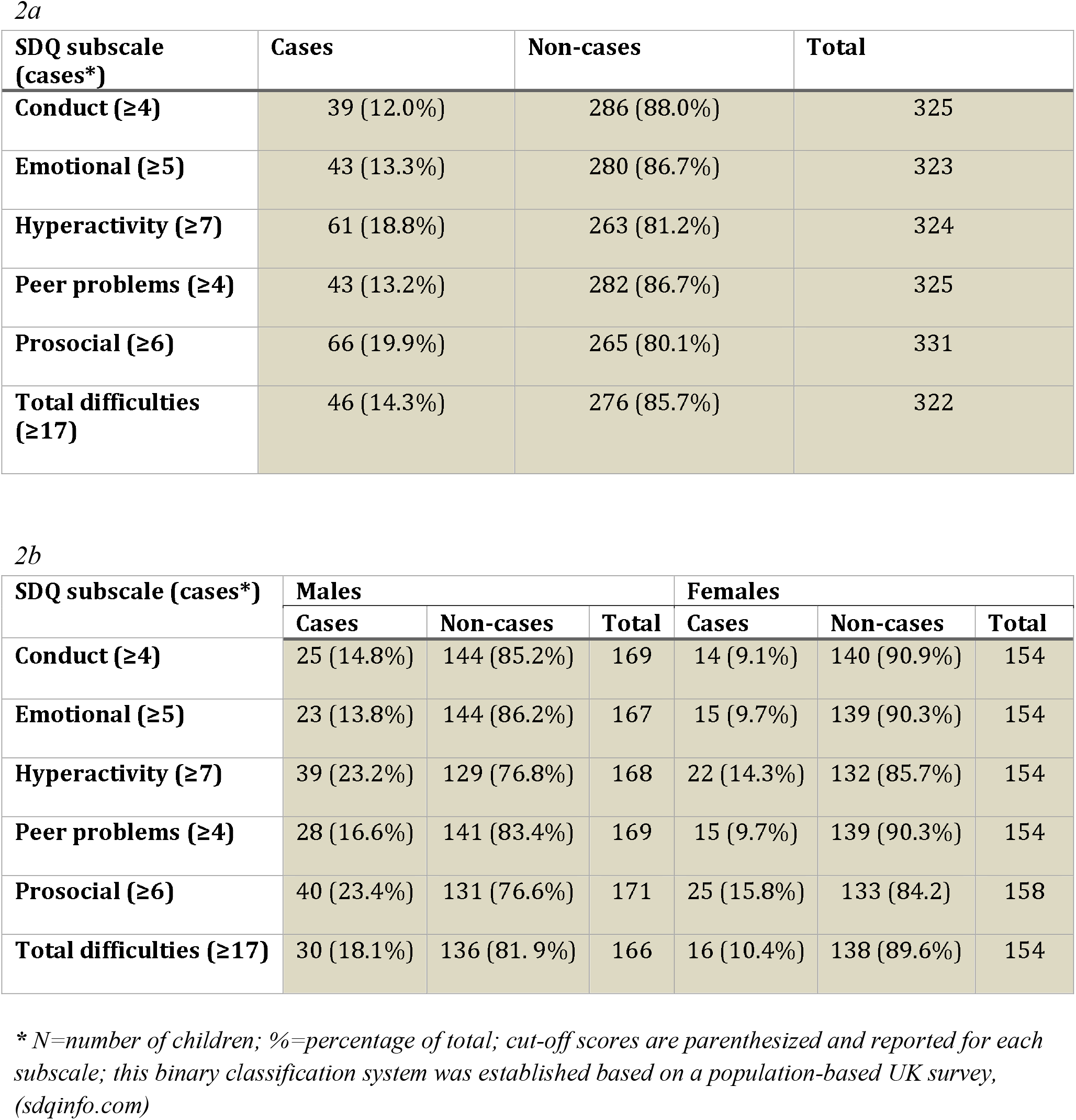
Prevalence of Behavioural Problems across SDQ subscales in children born with CL/P from the Cleft Collective. **Table 2a** presents the prevalence across the total sample and **2b** stratified by gender.

### Strengths and Difficulties Compared to the General Population

Table 3 shows mean SDQ scores and standard deviation (SD), and the frequency (n) and percentage (%) of cases and controls in the Cleft Collective and the three general population groups. Overall, the percentage of cases in the Cleft Collective (14.3%) was greater than the percentage within all three general population estimates for total difficulties (MCS, 7.5%; ONS, 9.8%; ALSPAC, 6.6%).

**Table 3:** Summary of SDQ Scores and Behavioural Problems in the Cleft Collective 5-year old Cohort Compared to General Population Estimates

| SDQ subscale (*cases\**) | Cohort | Total | Mean (SD) | Cases | Non-cases |
| --- | --- | --- | --- | --- | --- |
| **Conduct ($\geq 4$)** | Cleft Collective | 325 | 1.68 (1.71) | 39 (12.0%) | 286 (88.0%) |
|  | MCS | 12808 | 1.40 (23.95) | 1265 (9.9%) | 11543 (90.1%) |
|  | ONS norms | 10298 | 1.60 (1.70) | 1301 (12.6%) | 8997(87.4%) |
|  | ALSPAC | 9371 | 1.96 (1.40) | 1244 (13.0%) | 8127 (87.0%) |
| **Emotional ($\geq 5$)** | Cleft Collective | 323 | 1.93 (2.13) | 43 (13.3%) | 280 (86.7%) |
|  | MCS | 12780 | 1.30 (0.00) | 950 (7.4%) | 11830 (92.6%) |
|  | ONS norms | 10298 | 1.90 (2.00) | 1166 (11.3%) | 9132 (88.7%) |
|  | ALSPAC | 9385 | 1.44 (1.51) | 430 (4.6%) | 8955 (95.4%) |
| **Hyperactivity ($\geq 7$)** | Cleft Collective | 324 | 4.05 (2.79) | 61 (18.8%) | 263 (81.2%) |
|  | MCS | 12760 | 3.20 (0.00) | 1634 (12.8%) | 11126 (87.2%) |
|  | ONS norms | 10298 | 3.60 (2.70) | 1058 (14.6%) | 8790 (85.4%) |
|  | ALSPAC | 9377 | 3.39 (2.37) | 1342 (14.3%) | 8035 (85.7%) |
| **Peer problems ($\geq 4$)** | Cleft Collective | 325 | 1.45 (1.83) | 43 (13.2%) | 282 (86.7%) |
|  | MCS | 12786 | 1.00 (11.60) | 1161 (9.1%) | 11625 (90.9%) |
|  | ONS norms | 1215 | 1.40 (1.70) | 1215 (11.8%) | 9083 (88.2%) |
|  | ALSPAC | 9384 | 1.52 (1.48) | 1012 (10.8) | 8372 (89.2) |
| **Prosocial ($\geq 6$)** | Cleft Collective | 331 | 8.30 (1.87) | 66 (19.9%) | 265 (80.1%) |
|  | MCS | 12811 | 8.40 (0.00) | 1610 (12.6%) | 11201 (87.4%) |
|  | ONS norms | 10298 | 8.60 (1.60) | 1094 (10.6%) | 9204 (89.4%) |
|  | ALSPAC | 9372 | 7.04 (1.97) | 2207 (23.5%) | 7165 (76.5%) |
| **Total difficulties ($\geq 17$)** | Cleft Collective | 322 | 9.08 (6.45) | 46 (14.3%) | 276 (85.7%) |
|  | MCS | 12703 | 6.70 (11.99) | 947 (7.5%) | 11756 (92.5%) |
|  | ONS norms | 10298 | 8.60 (5.70) | 1009 (9.8%) | 9289 (90.2%) |
|  | ALSPAC | 9342 | 8.89 (4.56) | 614 (6.6%) | 8728 (93.4%) |

*\*Cut-off scores reported for each subscale; this binary classification system was established based on a population-based UK survey, (sdqinfo.com); SD=standard deviation; CC=Cleft Collective 5-year old cohort; MCS=Millennium Cohort study (CLS, 2008) represents the primary control group; ONS Norms=normative school-age SDQ data from Britain (Meltzer, 2000); ALSPAC= Avon Longitudinal Study of Parents and Children age 4 data*

Odds ratios (OR), 95% confidence intervals (CI), and p values comparing the relative odds of cases in the Cleft Collective to the three general population estimates are reported in Table 4. We found strong evidence to suggest that 5-year-olds in the Cleft Collective are more likely to experience behavioural problems than those in all three general population cohorts: MCS (OR = 2.07 [95% CI = 1.50-2.85]; P<.001), ONS Norms (OR = 1.53 [95% CI = 1.12-2.11]; P=.008), and ALSPAC (OR = 2.37 [95% CI = 1.72-3.27]; P<.001).

**Table 4:** Odds Ratios for Behavioural Problems in the Cleft Collective Compared to the General Population Samples.

| SDQ subscale | Cohort | Odds Ratio | 95%CI | p value |
| --- | --- | --- | --- | --- |
| Conduct ($\geq 4$) | Cleft Collective | 1.00 | | |
|  | MCS | 1.24 | (0.89, 1.75) | 0.207 |
|  | ONS norms | 0.94 | (0.67, 1.32) | 0.735 |
|  | ALSPAC | 0.89 | (0.63, 1.25) | 0.505 |
| Emotional ($\geq 5$) | Cleft Collective | 1.00 | | |
|  | MCS | 1.91 | (1.38, 2.65) | <0.001 |
|  | ONS norms | 1.20 | (0.87, 1.67) | 0.268 |
|  | ALSPAC | 3.20 | (2.29, 4.47) | <0.001 |
| Hyperactivity ($\geq 7$) | Cleft Collective | 1.00 | | |
|  | MCS | 1.58 | (1.19, 2.10) | 0.002 |
|  | ONS norms | 1.93 | (1.45, 2.56) | <0.001 |
|  | ALSPAC | 1.39 | (1.04, 1.85) | 0.024 |
| Peer problems ($\geq 4$) | Cleft Collective | 1.00 | | |
|  | MCS | 1.53 | (1.10, 2.12) | 0.011 |
|  | ONS norms | 1.14 | (0.82, 1.58) | 0.432 |
|  | ALSPAC | 1.26 | (0.91, 1.75) | 0.164 |
| Prosocial ($\geq 6$) | Cleft Collective | 1.00 | | |
|  | MCS | 1.73 | (1.32, 2.28) | <0.001 |
|  | ONS norms | 2.10 | (1.59, 2.76) | <0.001 |
|  | ALSPAC | 0.81 | (0.61, 1.06) | 0.128 |
| Total difficulties ($\geq 17$) | Cleft Collective | 1.00 | | |
|  | MCS | 2.07 | (1.50, 2.85) | <0.001 |
|  | ONS norms | 1.53 | (1.12, 2.11) | 0.008 |
|  | ALSPAC | 2.37 | (1.72, 3.27) | <0.001 |

*\*Cut-off scores reported for each subscale; this binary classification system was established based on a population-based UK survey, (sdqinfo.com); 95% CI=95% Confidence Intervals; CC=Cleft Collective 5-year old cohort; MCS=Millennium Cohort study (CLS, 2008) represents the primary control group; ONS Norms=normative school-age SDQ data from Britain (Meltzer, 2000); ALSPAC= Avon Longitudinal Study of Parents and Children age 4 data*

Specifically, hyperactivity, emotional, and peer problems were more prevalent in the Cleft Collective than in all three general population estimates, and prosocial problems more prevalent than in two of the three cohorts (Table 3). There was strong evidence of increased odds of hyperactivity among those in the Cleft Collective compared with individuals from each of the population cohorts (Table 3). For example, individuals in the cleft group were almost two times more likely to experience hyperactivity compared with individuals in the ONS norms group (OR 1.93, 95%CI 1.45, 2.56, p <0.001). Furthermore, we found strong evidence suggesting greater odds of emotional problems among children in the Cleft Collective compared with MCS (OR = 1.91 [95% CI = 1.38-2.65]; P<.001) and ALSPAC (OR = 3.20 [95% CI = 2.29-4.47]; P<.001).

Gender stratified analyses indicated that emotional problems were particularly prevalent among boys (Supplement 1), with boys in the Cleft Collective nearly twice as likely as those in the MCS (OR = 1.92 [95% CI = 1.23-3.01]; P=.004) and three times those in ALSPAC (OR = 3.31 [95% CI = 2.09-5.25]; P<.001) to experience emotional difficulties (Supplement 2). Gender-stratified analyses are reported in Supplement 1 and Supplement 2.

### Maternal, familial and cleft-related factors associated with behavioural problems in the Cleft Collective

Among mother’s who completed the SDQ response rates on questions informing predictors were high in all cases (≥ 91.5%), with the exceptions of cleft type (n=170 (50%)), smoking status (n=157 (46%)) and syndrome (n=94 (27%)) (Table 1).

Figure 1 illustrates associations between maternal and familial variables and the odds of behavioural problems in the Cleft Collective, reporting odds ratios, gender-adjusted odds ratios, and 95% confidence intervals (CI) from a series of logistic regression analyses.

**Figure 1:**
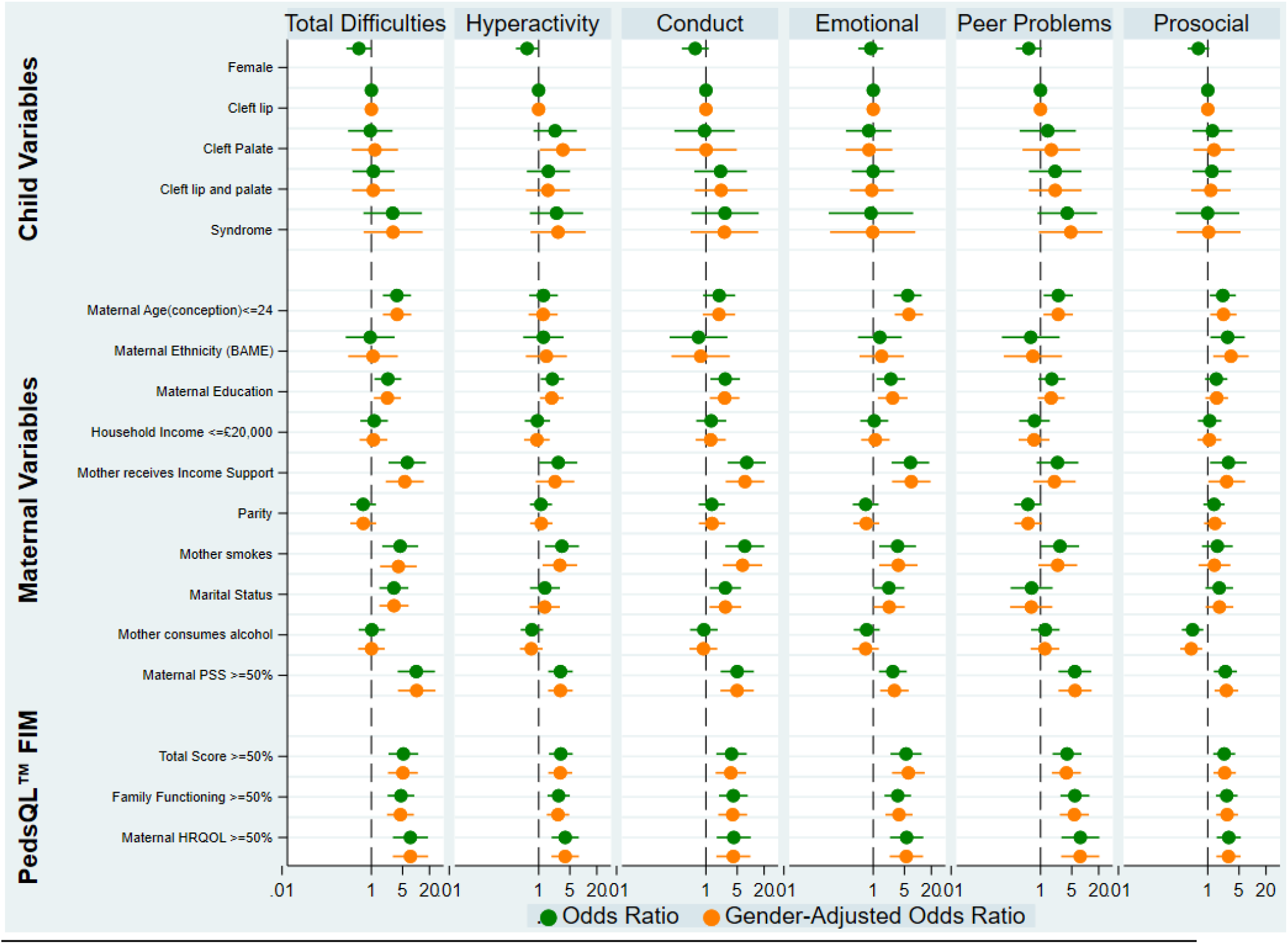
Associations between Sociodemographic and Maternal Characteristics and Behavioural problems among CL/P 5-Year-Olds *This figure shows the results of logistic regression analyses between thirteen predictor variables and cases; this binary classification system was established based on a population-based UK survey, (sdqinfo.com); Odds ratios and gender-adjusted odds ratios with 95% confidence intervals are reported across SDQ subscales;

High maternal psychological stress, as indicated by scores above the 50^th^ percentile on the Perceived Stress Scale (PSS), showed the strongest association, with higher odds of behavioral difficulties in children (OR=10.1, 95% CI=3.87-26.51). High maternal and family psychological impact, as indicated by scores above the 50^th^ percentile on the Pediatric Quality of Life Inventory Family Impact Module (PedsQL™ FIM), also showed strong evidence of association with higher odds of behavioral difficulties in children (OR=5.20, 95% CI=2.42-11.19). The odds of behavioural problems in children whose families received income support was 6-fold that of those whose families did not (OR=6.35, 95% CI=2.42-16.68). However, there was very little evidence of association of household income (<=£20,000 versus >£20,000) with the odds of behavioural problems (OR=1.2, 95% CI=0.57-2.31).

There was strong evidence of an association between the mother not being married OR=3.19, 95% CI=1.51-6.72), being a smoker (OR=4.41, 95% CI=1.75-11.13), maternal age at conception younger than 24 years (OR=3.73, 95% CI=1.81-7.72) and higher odds of behavioral difficulties in children born with CL/P. There was also good evidence of an association between lower maternal education (mother not having a higher education degree) (OR=2.34, 95% CI=1.17-4.69) and higher odds of behavioural problems in the study cohort (Figure 1).

Maternal ethnicity did not show evidence of association with the odds of behavioural problems in children, as defined by total difficulties score and across subscales. However there was evidence of higher odds of prosocial difficulties among children whose mothers self-reported as Black, Asian, or of another minority ethnic group in the UK (BAME) (Prosocial OR=2.78, 95% CI=1.14-6.74).

There was some evidence that children whose mothers drank alcohol had higher odds of prosocial behavior (OR=0.46, 95% CI=0.26-0.79), and having more than one child was associated with lower odds of peer problems, when estimates were adjusted for gender (OR (gender adjusted) =0.47, 95% CI=0.23-0.96).

Although there was moderate evidence to suggest the odds of behavioural problems in girls were lower than for boys (OR=0.53, CI=0.27-1.01), gender had a negligible effect on the other predictor variables explored, as represented by the closely aligned gender-adjusted odds ratios (Figure 1).

## Discussion

### Key Findings

In this study, we reported that an estimated 14.3% of 5-year-old children with CL/P experience behavioural problems as measured by the SDQ total difficulties score. Findings suggested that children born with a cleft experience more behavioural problems, and specifically more hyperactivity-and emotional-related difficulties compared to the general population. Greater peer problems, and decreased prosocial behaviors were also more prevalent in the Cleft Collective relative to the general population, suggesting unmet needs in these areas.

Our findings also showed that known maternal and familial factors which are associated with increased behavioural problems in the general population, were also associated with behavioural problems in children born with CL/P. Mainly, lower maternal education, lower income, lower maternal well-being, income, lower family functioning and marital status, showed associations with increased behavioural problems within the cohort.

### Comparison with Other Studies

Our findings support previous research findings on the prevalence of behavioural problems in children born with cleft. Specifically, our study is in agreement with previous investigations from Cleft Care UK study which reported increased hyperactivity-related difficulties (measured by parent-reported SDQ) in 5-year old children born with unilateral cleft lip and palate (Waylen et al., 2017). Increased hyperactivity-related difficulties, but not other behavioural problems, were found in 6-12 year old children born with cleft from the United States (Wehby et al., 2012). In addition, a recent longitudinal study reported increased behavioral difficulties, as measured by the SDQ at ages 5 and 10 years, with boys born with cleft reporting more difficulties (Pinckston et al., 2020). Emotional difficulties and deficient social competencies have also been found in school-aged children born with cleft (Murray et al., 2010; Waylen et al., 2017). However, increased behavioral difficulties in children born with cleft were not reported in other studies, including a cross-sectional study of 11-16 year old children born with cleft (Berger & Dalton, 2009).

Maternal and familial factors, including maternal smoking, mother not being married, younger maternal age at conception, lower maternal education, receiving income support, high maternal psychological stress and high maternal and family psychological impact, showed the strongest associations with behavioural problems in 5-year-olds born with CL/P. Maternal ethnicity did not appear to be associated with behavioural problems (as defined by total difficulties) and across subscales, however there was evidence that maternal ethnicity was associated with increased prosocial difficulties. For consistency with MCS findings (Borrell-Porta M, 2017) and due to small sample sizes, we explored ethnicity as a binary variable, and grouped mothers in the cohort who identify as Black, Asian, or as belonging to a minority ethnic group in the UK. We recognize that this is a major limitation of our exploration of ethnicity and that by doing this we are unable to draw conclusions about the unique needs and experiences of individuals from different ethnic groups.

One of the strongest associations we found was between high maternal perceived psychological stress (PSS), reported when children were 5 years old and behavioural problems at age 5. Previous studies have shown evidence of strong associations between maternal stress and child behavioural problems in both the general population and children born with cleft (M. C. Endriga et al., 2003; M. C. Endriga & Kapp-Simon, 1999; Kovess-Masfety et al., 2016; Stock, Costa, White, & Rumsey, 2020; Thamilselvan et al., 2015). Whether this association is stronger in children born with cleft warrants further investigation (Thamilselvan et al., 2015). In addition, we were not able to establish causality and direction of effects as explained in the limitations section.

Family functioning has been linked to child psychological health and is known to be a particularly strong determinant of well-being in children born with chronic medical conditions (Macho, Bohac, Fedeles, Fekiacova, & Fedeles, 2017; J. W. Varni, Seid, & Rode, 1999). Our findings provided evidence of this association in children born with cleft, demonstrating the strong association between behavioural problems and measures of family functioning. Though the merits of the PedsQL™ FIM for measuring the impact of pediatric chronic health conditions has been well-established (Medrano, Berlin, & Hobart Davies, 2013; Mishra, Ramachandran, Firdaus, & Rath, 2015; J. W. Varni et al., 1999; James W. Varni et al., 2004), the tool is relatively new to the cleft literature, and our study joins few others (Stock et al., 2020). In one recent study using the PedsQL™ FIM, family functioning was reported to be higher by (n=1,163 parents) following a diagnosis of CL/P, compared to normative data (Stock et al., 2020). Our results reinforce the utility of this tool in children born with cleft (Crerand et al., 2015).

### Strengths and Limitations

There are many key strengths to this study. Firstly, the sample size is large, geographically diverse across the UK, and age specific. Secondly, UK cleft research is supported by standard provision and quality of patient care, relative to other countries. This homogeneity, augmented by the centralization of cleft care over the last two decades, offers major benefits for comparing patient outcomes and for evaluating the effectiveness of interventions implemented (Ness et al., 2015). Thirdly, three UK population cohorts of similar age to Cleft Collective were used as controls. Different biases and confounding structures might have been present in each cohort depending on recruitment and data collection procedures and the fact that our results were in the same direction across all cohorts strengthens our confidence in them.

The limitations of our study are inherent to its observational nature. Aside from true causal effects of cleft and maternal and familial variables on behavioral difficulties, the reported associations could be explained by bias or confounding. For example, reverse causation could be at play in the association between maternal perceived stress and behavioural problems in cleft. Having a child with behavioural problems could be causing increased psychological stress in the parents and not the other way around. Another possibility is that mothers with high levels of stress could be reporting more behavioral problems in their children than mothers with lower levels of stress. The same could be true for family functioning. Other associations could be confounded by several measured and unmeasured factors which we were not able to control for. Alternative research designs are needed to infer causation and Mendelian randomization (MR) offers one approach by reducing bias from confounding and reverse causation using observational data (Lawlor, Tilling, & Davey Smith, 2016).

Although we observed more behavioural problems among children with syndromic cleft versus those with non-syndromic cleft, we were unable to assess the prevalence of behavioural problems by cleft type and compare children with syndromic cleft versus those with non-syndromic cleft due to small sample sizes (n<10).

In our study we did not investigate clinical diagnoses of conditions, such as attention deficit/hyperactivity disorder (ADHD), autism, dyslexia, specific language impairment (SLI), and developmental delay, which could be overrepresented in cleft populations (Feragen, Stock, & Rumsey, 2014). However, high scores on the SDQ have been found to be strongly linked to clinical diagnoses of childhood psychiatric disorders, such as ADHD (Russell, Rodgers, & Ford, 2013). In addition, in 5-year old children some of these conditions, such as ADHD, are unlikely to have been diagnosed and following participants from the Cleft Collective at older ages would be required to investigate clinical diagnoses of childhood psychiatric disorders in cleft.

Another limitation linked to the observational nature of the study is missingness. Our analyses were performed on children whose mothers returned the baseline questionnaire at 5 years of age and completed the SDQ section. However, other parts of the questionnaire were sometimes missing and this reduced our power to detect associations with maternal, familial and cleft-related factors. In addition, missingness might not have been random; e.g. parents might have been less likely to answer the smoking questions if they had smoked during pregnancy.

Despite this limitation, it is reassuring to detect evidence of association of many of the factors that have been associated with behavioural problems in the general population.

Behavioral difficulties in cleft might differ according to syndromic or non-syndromic status and cleft type. Due to sample size limitations, we were not powered to perform stratified analyses and we aim to follow our findings in a larger sample size as recruitment to the Cleft Collective in still ongoing.

A final concern is the study’s reliance on a single informant (mothers) to provide information on behavioural problems. Many studies delineate the value of using a multi-informant approach to assess a child’s mental health, highlighting the improved sensitivity and specificity for detecting disorders that comes with this. However, the psychometric properties of the parent version of the SDQ are strong in 5-year-olds (Mieloo et al., 2012; Stone et al., 2010), and have been further confirmed in cleft studies (Bjerke, Feragen, & Bergvik, 2018).

### Implications for policy and practice

Our findings suggest that there are health disparities among the CL/P population and that many predictors of behavioral difficulties in the general population, such as income and maternal education, can be extended to the CL/P population. There is a need for targeted interventions which can provide early support to children born with CL/P who are at an elevated risk of poor mental health outcomes. Our results support the need for integrated psychological support (i.e. the presence of mental health specialists), on cleft teams and reinforce the imperative for routine psychological assessments of children born with cleft (O. Hunt, Burden, Hepper, Stevenson, & Johnston, 2006; “Improving support for people with cleft lip and palate -UWE Bristol: Research with impact.,”).

## Conclusion

We found that children born with cleft, particularly boys with CL/P, experience more behavioural problems as indicated by SDQ total difficulties scores, and specifically more hyperactivity-and emotional-related difficulties compared to the general population. This study is limited in its ability to make causal inferences about mental health challenges in children born with cleft, and leaves room for future inquiry into the direction of the associations observed (Lawlor et al., 2016). Further investigation is also necessary to clarify the extent and severity of difficulties in high-risk cleft populations, and to unravel the pathways and mechanisms driving outcomes.

## Data Availability

Data are available from the Cleft Collective after submitting a research proposal.

## Acknowledgements

This publication involves data derived from independent research funded by The Scar Free Foundation (REC approval 13/SW/0064). We are grateful to the families who participated in the study, the UK NHS cleft teams, and The Cleft Collective team, who helped facilitate the study. The views expressed in this publication are those of the authors and not necessarily those of The Scar Free Foundation or The Cleft Collective Cohort Studies team. We are extremely grateful to all the families who took part in the ALSPAC study, the midwives for their help in recruiting them, and the whole ALSPAC team, which includes interviewers, computer and laboratory technicians, clerical workers, research scientists, volunteers, managers, receptionists and nurses. We are grateful to the Centre for Longitudinal Studies (CLS), UCL Social Research Institute, for the use of the Millennium Cohort Study data and to the UK Data Service for making them available. However, neither CLS nor the UK Data Service bear any responsibility for the analysis or interpretation of these data. I acknowledge the University of Bristol and the MSc Public Health program.

## Funding

The authors disclosed receipt of the following financial support for the research, authorship, and/or publication of this article: Sammy Berman was supported by the US-UK Fulbright Commission and IIE [University of Bristol Partnership Award]; and Foundation for Faces of Children [Dr. John Mulliken Award]. This study was supported by grant 204895/Z/16/Z from the Wellcome Trust (Rachel Blakey), which was awarded to Anita Thapar, Kate Tilling, Evie Stergiakouli, and George Davey Smith. For the purpose of Open Access, the author has applied a CC BY public copyright license to any Author Accepted Manuscript version arising from this submission. The Medical Research Council (MRC) and the University of Bristol support the MRC Integrative Epidemiology Unit (grants MC_UU_00011/1 and MC_UU_00011/3). The UK Medical Research Council and Wellcome (Grant ref: 217065/Z/19/Z) and the University of Bristol provide core support for ALSPAC. GCS is financially supported by the Medical Research Council [New Investigator Research Grant, MR/S009310/1] and the European Joint Programming Initiative “A Healthy Diet for a Healthy Life” (JPI HDHL, NutriPROGRAM project, UK MRC MR/S036520/1].

This publication is the work of the authors and Dr Evie Stergiakouli will serve as guarantor for the contents of this paper.

