## Supplemental Tables for "Prevalence and Factors Associated with Behavioural Problems in 5-year-old Children Born with Cleft Lip and/or Palate from the Cleft Collective"

**Supplement 1: Summary of SDQ Scores and Behavioural Problems in the Cleft Collective 5-year old Cohort Compared to General Population Estimates Stratified by Gender**

| **SDQ subscale** | **Cohort** | **Total males** | **Male cases** | **Male non-cases** | **Total Females** | **Female**  **cases** | **Female**  **Non-cases** |
| --- | --- | --- | --- | --- | --- | --- | --- |
| Conduct (≥4) | Cleft Collective | 169 | 25 (14.8%) | 144 (85.2%) | 154 | 14 (9.1%) | 140 (90.9%) |
|  | MCS | 6501 | 789 (12.1%) | 5712 (87.9%) | 6307 | 476 (7.5%) | 5831 (92.5%) |
|  | ONS norms | 5153 | 778 (15.1% | 4375 (84.9%) | 5145 | 531 (10.3%) | 4614 (89.7%) |
|  | ALSPAC | 4849 | 724 (14.9%) | 4123 (85.0%) | 4521 | 520 (11.5%) | 4001 (88.5%) |
| Emotional (≥5) | Cleft Collective | 167 | 23 (13.8%) | 144 (86.2%) | 154 | 15 (9.7%) | 139 (90.3%) |
|  | MCS | 6488 | 498 (7.7%) | 5990  (92.3%) | 6292 | 452 (7.2%) | 5840  (92.8%) |
|  | ONS norms | 5153 | 548 (10.6%) | 4605 (89.4%) | 5145 | 623 (12.1%) | 4522 (87.9%) |
|  | ALSPAC | 4829 | 223 (4.6%) | 4626 (95.8%) | 4533 | 207 (4.6) | 4326 (95.4%) |
| Hyperactivity (≥7) | Cleft Collective | 168 | 39 (23.2%) | 129 (76.8%) | 154 | 22 (14.3%) | 132 (85.7%) |
|  | MCS | 6480 | 1069 (16.5%) | 5411 (83.5%) | 6280 | 565 (9.0%) | 5715 (91.0%) |
|  | ONS norms | 5153 | 1005 (19.5%) | 4148 (80.5%) | 5145 | 504 (9.8%) | 4641 (9.1%) |
|  | ALSPAC | 4843 | 828 (17.1) | 4015 (82.9) | 4531 | 514 (11.3%) | 4017 (88.7%) |
| Peer problems (≥4) | Cleft Collective | 169 | 28 (16.6%) | 141 (83.4%) | 154 | 15 (9.7%) | 139 (90.3%) |
|  | MCS | 6491 | 692 (10.7%) | 5799 (89.3%) | 6295 | 469 (7.5%) | 5826 (92.5%) |
|  | ONS norms | 5153 | 687 (13.3%) | 4466 (86.7%) | 5145 | 520 (10.1%) | 4625 (89.9%) |
|  | ALSPAC | 4850 | 592 (12.2%) | 4258 (87.8%) | 4531 | 420 (9.3%) | 4111 (90.7%) |
| Prosocial (≥6) | Cleft Collective | 171 | 40 (23.4%) | 131 (76.6%) | 158 | 25 (15.8%) | 133 (84.2) |
|  | MCS | 6504 | 1084 (16.7%) | 5420 (83.3%) | 6307 | 526 (8.3%) | 5781 (91.7%) |
|  | ONS norms | 5153 | 720 (14.0%) | 4433 (86.0%) | 5145 | 364 (7.1%) | 4781 (92.9%) |
|  | ALSPAC | 4846 | 1382 (28.5%) | 3464 (71.5%) | 4523 | 824 (18.2%) | 3699 (81.8%) |
| Total difficulties (≥17) | Cleft Collective | 166 | 30 (18.1%) | 136 (81. 9%) | 154 | 16 (10.4%) | 138 (89.6%) |
|  | MCS | 6449 | 610 (9.5%) | 5839 (90.5%) | 6254 | 337 (5.4%) | 5917 (94.6%) |
|  | ONS norms | 5153 | 614 (11.9%) | 4539 (88.3%) | 5145 | 401 (7.8%) | 4745 (91.9%) |
|  | ALSPAC | 4829 | 388 (8%) | 4441 (92.0%) | 4510 | 226 (5.0%) | 4284 (95.0%) |

**Cut-off scores reported for each subscale; this binary classification system was established based on a population-based UK survey, (sdqinfo.com)*; *CC=Cleft Collective 5-year old cohort; MC=Millennium Cohort study (CLS, 2008) represents the primary control group; ONS Norms=normative school-age SDQ data from Britain (Meltzer, 2000); ALSPAC= Avon Longitudinal Study of Parents and Children age 4 data*

**Supplement 2: Odds Ratios for Behavioural Problems in the Cleft Collective Compared to the General Population Samples Stratified by Gender**

| SDQ subscale | Cohort | Male Only | | | Female Only | | |
| --- | --- | --- | --- | --- | --- | --- | --- |
|  |  | Odds Ratio | 95% CI | p value | Odds Ratio | 95% CI | p value |
| Conduct (≥4) | Cleft Collective | 1 |  |  | 1.00 |  |  |
|  | MCS | 1.26 | (0.82, 1.93) | 0.299 | 1.23 | (0.70, 2.14) | 0.475 |
|  | ONS norms | 0.98 | (0.63, 1.50) | 0.913 | 0.87 | (0.50, 1.52) | 0.621 |
|  | ALSPAC | 0.99 | (0.64, 1.52) | 0.959 | 0.77 | (0.44, 1.34) | 0.357 |
| Emotional (≥5) | Cleft Collective | 1.00 |  |  | 1.00 |  |  |
|  | MCS | 1.92 | (1.23, 3.01) | 0.004 | 1.39 | (0.81, 2.40) | 0.229 |
|  | ONS norms | 1.34 | (0.86, 2.10) | 0.199 | 0.78 | (0.46, 1.34) | 0.375 |
|  | ALSPAC | 3.31 | (2.09, 5.25) | <0.001 | 2.26 | (1.30, 3.91) | 0.004 |
| Hyperactivity (≥7) | Cleft Collective | 1.00 |  |  | 1.00 |  |  |
|  | MCS | 1.53 | (1.06, 2.20) | 0.022 | 1.69 | (1.06, 2.67) | 0.026 |
|  | ONS norms | 1.25 | (0.87, 1.80) | 0.234 | 1.53 | (0.97, 2.43) | 0.068 |
|  | ALSPAC | 1.47 | (1.02, 2.11) | 0.040 | 1.30 | (0.82, 2.06) | 0.261 |
| Peer problems (≥4) | Cleft Collective | 1.00 |  |  | 1.00 |  |  |
|  | MCS | 1.66 | (1.10, 2.52) | 0.016 | 1.34 | (0.78, 2.30) | 0.288 |
|  | ONS norms | 1.29 | (0.85, 1.95) | 0.226 | 0.96 | (0.56, 1.65) | 0.882 |
|  | ALSPAC | 1.43 | (0.94, 2.16) | 0.092 | 1.06 | (0.61, 1.82) | 0.843 |
| Prosocial(≥6) | Cleft Collective | 1.00 |  |  | 1.00 |  |  |
|  | MCS | 1.53 | (1.07, 2.19) | 0.021 | 2.07 | (1.34, 3.20) | <0.001 |
|  | ONS norms | 1.88 | (1.31, 2.70) | <0.001 | 2.47 | (1.59, 3.83) | <0.001 |
|  | ALSPAC | 0.77 | (0.53, 1.10) | 0.145 | 0.84 | (0.55, 1.30) | 0.358 |
| Total difficulties (≥17) | Cleft Collective | 1.00 |  |  | 1.00 |  |  |
|  | MCS | 2.11 | (1.41, 3.16) | <0.001 | 2.04 | (1.20, 3.46) | 0.008 |
|  | ONS norms | 1.63 | (1.09, 2.44) | 0.018 | 1.37 | (0.81, 2.33) | 0.592 |
|  | ALSPAC | 2.52 | (1.68, 3.80) | <0.001 | 2.20 | (1.29, 3.75) | <0.001 |

**Cut-off scores reported for each subscale; this binary classification system was established based on a population-based UK survey, (sdqinfo.com)*; *95% CI=95% Confidence Intervals; CC=Cleft Collective 5-year old cohort; MCS=Millennium Cohort study (CLS, 2008) represents the primary control group; ONS Norms=normative school-age SDQ data from Britain (Meltzer, 2000); ALSPAC= Avon Longitudinal Study of Parents and Children age 4 data*
